# Cardiac magnetic resonance markers of pre-clinical hypertrophic and dilated cardiomyopathy in genetic variant carriers

**DOI:** 10.1101/2024.11.15.24317368

**Authors:** Philip M. Croon, Marion van Vugt, Cornelis P. Allaart, Bram Ruijsink, Perry M. Elliott, Folkert W. Asselbergs, Rohan Khera, Connie R. Bezzina, Michiel Winter, A. Floriaan Schmidt

**Affiliations:** Department of Cardiology, Amsterdam Cardiovascular Sciences, Amsterdam University Medical Centre, University of Amsterdam, Amsterdam, The Netherlands; Section of Cardiovascular Medicine, Department of Internal Medicine, Yale School of Medicine, New Haven, CT; Amsterdam Cardiovascular Sciences, Heart Failure and Arrhythmias, Amsterdam, The Netherlands; Division Heart & Lungs, Department of Cardiology, University Medical Center Utrecht, Utrecht University, Utrecht, The Netherlands; Institute of Cardiovascular Science, Faculty of Population Health, University College London, London, United Kingdom; School of Biomedical Engineering & Imaging Sciences, King’s College London, London, United Kingdom; Barts Heart Centre, St. Bartholomew’s Hospital, London, United Kingdom; Centre for Heart Muscle Disease, Institute of Cardiovascular Science, University College London, London, United Kingdom; Institute of Health Informatics, University College London, London, UK; National Institute for Health Research, University College London Hospitals, Biomedical Research Centre, University College London, London, UK; Section of Health Informatics, Department of Biostatistics, Yale School of Public Health, New Haven, CT; Section of Biomedical Informatics and Data Science, Yale School of Medicine, New Haven, CT; Center for Outcomes Research and Evaluation, Yale-New Haven Hospital, New Haven, CT; Department of Experimental Cardiology, Amsterdam Cardiovascular Sciences, Amsterdam University Medical Centre, University of Amsterdam, Amsterdam, The Netherlands; UCL British Heart Foundation Research Accelerator, London, United Kingdom

**Keywords:** Dilated cardiomyopathy, cardiac magnetic resonance, hypertrophic cardiomyopathy, whole exome sequencing, genetics

## Abstract

**Background:** Patients with hypertrophic cardiomyopathy (HCM) and dilated cardiomyopathy (DCM) exhibit structural and functional cardiac abnormalities. We aimed to identify imaging biomarkers for pre-clinical cardiomyopathy in healthy individuals carrying cardiomyopathy-associated variants (G+).

**Methods:** We included 40,169 UK biobank participants without cardiac disease who had cardiac magnetic resonance imaging (CMR) measurements and whole exome sequencing. We first validated 22 CMR measurements by associating them with incident atrial fibrillation (AF) or heart failure (HF). We subsequently assessed associations of these measurements with HCM or DCM G+, or specific genes, utilising generalised linear models conditional on cardiac risk factors.

**Results:** Thirteen CMR measurements were associated with incident AF and fifteen with HF. These included left-ventricular (LV) ejection fraction (EF) (hazard ratio [HR] 0.61, 95% confidence interval [95%CI] 0.54; 0.69) for HF and indexed maximum left atrial volume (LA-Vi max; HR1.47, 95%CI 1.29; 1.67) for AF. Five measurements associated with HCM G+, amongst which right ventricular (RV) end-systolic volume (RV-ESV; OR 0.62, 95%CI 0.53; 0.74), RV-EF (OR 1.36, 95%CI 1.19; 1.55), and right atrial EF (OR 1.22, 95%CI 1.08; 1.39). All associations overlapping with incident disease and HCM had opposite effect directions, such as RV-ESV with HF (OR 1.22, 95%CI 1.07; 1.40). Two CMR measurements associated with DCM G+: LV-ESVi (OR 1.35, 95%CI 1.15; 1.58) and LV-EF (OR 0.75, 95%CI 0.64; 0.88). Due to heterogeneity, we explored associations with individual cardiomyopathy genes, finding MAPSE associated with *TTN* and *TNNT2*, and LA pump and RA-EF associated with *MYH7*.

**Conclusion:** We identified right heart CMR measurements associated with HCM G+ in healthy individuals, indicating early compensation of cardiac function. LV measurements were associated with DCM G+, where the CMR associations varied across individual DCM genes, suggesting distinct early pathophysiology.

## Background

Cardiomyopathies are a group of disorders characterised by structural abnormalities of the heart which may lead to impaired cardiac function^1^. These abnormalities often contribute to the development of conditions such as heart failure (HF), atrial fibrillation (AF), and can lead to sudden cardiac death^2,3^. Hypertrophic cardiomyopathy (HCM) and dilated cardiomyopathy (DCM) are the two most common cardiomyopathies^4^. HCM is characterised by ventricular hypertrophy unexplained by abnormal loading conditions^2^, whereas DCM is characterised by dilatation and impaired systolic function of the left ventricle or both ventricles^4^.

Cardiomyopathies often have strong genetic aetiology, with pathogenic genetic variants found in up to 40% of HCM and DCM patients^1,5,6^. The prevalence of pathogenic variants in the general population depends on the variant adjudication, but has been estimated to be up to 1 in 150 for HCM and 1 in 250 people for DCM^7^. Disease onset is extremely variable across carriers of a cardiomyopathy (CMP) variant. In some, pathogenic variants lead to childhood disease, while others are affected late in life or not at all. The understanding of the early cardiac phenotype in individuals carrying a pathogenic genetic variant and are free of cardiac disease is currently limited^2,5^.

Cardiac magnetic resonance imaging (CMR) serves as the gold standard for non-invasive quantification of cardiac structure and function, and hence is the cornerstone in the routine monitoring of individuals carrying CMP-associated variants (G+) and patients^1^. A relatively small number of CMR measurements, such as left ventricular (LV) ejection fraction (EF), end diastolic volume (EDV) and LV wall thickness, are currently used in clinical care to monitor patients.

With the advent of deep learning (DL), a larger number of additional measurements can now be derived automatically and reproducibly from standard CMR, which may lead to novel insights into markers of disease onset and progression^8^. Using genome-wide associations studies (GWAS), we and others have found that variants near CMP-causing genes, such as *TTN* and *BAG3*, associate with CMR measurements derived from healthy participants^9–11^. While the genetic associations suggest that CMR may contain valuable information for individuals carrying CMP-associated variants, GWAS exclude the rare causal CMP variants, and hence do not provide information on the association between CMR and G+.

In the current study, we leveraged data from 40,169 UK biobank participants with available CMR data. We applied a DL-based automatic CMR analysis to derive 22 CMR measurements^8^ and examined their association with participants carrying CMP-associated variants, as identified using whole exome sequencing (WES). First, we empirically validated the 22 derived CMR measurements by assessing their association with incident AF and HF. Next, we identified the subset of CMR measurements associating with any CMP G+, as well as the CMR measurements associating with the three most frequent genes for HCM and DCM. Finally, we cross-referenced these findings with previously identified common genetic CMP variants associating with CMR measurements.

## Methods

### Cardiomyopathy G+

G+ were identified using the WES data available for 469,779 participants from the UK Biobank. Pathogenic and likely pathogenic variants for HCM and DCM were identified, as described by Bourfiss *et al*.^7^. Briefly, we selected genes classified to have definite, strong or moderate evidence of pathogenicity as defined by Clinical Genome Resource (ClinGen)^12^ and that were curated by Ingles *et al*.^13^ and Jordan *et al*.^14^. Variants in the following genes were identified for HCM: *ACTC1*, *CSRP3*, *JPH2*, *MYBPC3*, *MYH7*, *MYL2*, *MYL3*, *TNNC1*, *TNNI3*, *TNNT2,* and *TPM1*, and for DCM we included *ACTC1*, *ACTN2*, *BAG3*, *DES*, *DSP*, *FLNC*, *JPH2*, *LMNA*, *MYH7*, *NEXN*, *PLN*, *RBM20*, *SCN5A*, *TNNC1*, *TNNI3*, *TNNT2*, *TPM1*, *TTN*, and *VCL* (**Supplementary Table S1**). Next, likely pathogenic and pathogenic variants in these genes were identified using the ClinVar NCBI-NIH database and the Dutch Society for Clinical Genetic Laboratory Diagnostics (Vereniging Klinische Genetische Laboratoriumdiagnostiek) database. Some variants were associated with both HCM and DCM. Participants carrying these variants were included in both groups, as removing these participants did not affect the associations with clinical outcomes^7^.

To account for conflicting submissions in ClinVar, we performed a sensitivity analysis by including only the genetic variants with consistent pathogenic classification across all submissions.

### UK biobank participants

We utilized data from UK Biobank participants who enrolled to the CMR sub-study (n = 52,630). To account for a potential lag in registration or de novo diagnoses due to CMR, we excluded participants diagnosed with AF, HF, myocardial infarction, valvular disease, HCM, DCM, or CMP up to 30 days after the CMR (n = 2,888). The **Supplemental materials** detail the full data engineering strategy and **Supplementary Table S2** lists the employed diagnosis codes. Ethics approval for the UKB study was obtained from the North West Centre for Research Ethics Committee (11/NW/0382) and all participants provided informed consent.

### CMR measurements derivation

The full UK Biobank CMR protocol has been described in detail^15^. In short, all CMR examinations were performed on a 1.5 Tesla scanner (Magnetom Aera, Syngo Platform VD13A, Siemens Healthcare, Erlangen, Germany). We used a previously developed and validated DL model (AI-CMRQC) to extract 22 LV, right ventricular (RV), left (LA) or right atrial (RA) CMR measurement^8^). CMRQC contains the following steps:

1. The quality of the CMR is evaluated and artefacts are rejected.
2. The full cycle of both short axis, 4– and 2-chamber is segmented by a 17-layer 2D fully convolutional network.
3. LV and RV volume curves and LV mass were calculated, from which cardiac function parameters including end-diastolic (EDV) and end-systolic volumes (ESV), stroke volume, and EF were derived.
4. In the output, quality control profiles of volume curves and LV/RV consistency were evaluated by support vector machine classifiers and inconsistencies were rejected or revised.

Cine images of short-axis and 2– and 4-chamber long axis views were used to automatically calculate LV, RV, LA and RA functional measurements, including biventricular EDV, ESV, stroke volume (SV), EF, LA/RA minimal and maximal volume (V min and Vmax), EF, 2– and 4-chamber mitral annular plane systolic excursion (MAPSE 2/4Ch), and 2– and 4-chamber tricuspid annular plane systolic excursion (TAPSE 2/4Ch). Several CMR measurements were indexed (i) for body surface area.

### Statistical analysis

To validate the CMR protocol, the association between CMR and the onset of AF and HF were evaluated using a Cox semi-parametric proportional hazards model. Participants without established disease at baseline were followed from their CMR appointment until the earliest of the following events: onset of AF or HF separate, lost to follow-up, death, or administrative censoring, with a maximum follow-up of 6.5 years. These models were adjusted for age (in years), male sex, hypertension, diabetes, smoking (ever/never), and hypercholesterolemia. The model assumptions were evaluated by correlation of Schoenfeld residuals against time. Kaplan-Meier estimates of the cumulative AF or HF incidence were calculated by categorizing CMR measurements into an 85% “reference” group and a 15% “risk increasing” group.

The marginal associations between CMR measurements and G+ were ascertained using a generalised linear model with a binomial distribution and logit link function. Age and sex were included as covariates in a second model, with a third model additionally including cardiac risk factors: hypertension, diabetes, smoking status, and hypercholesterolaemia.

We additionally explored potential differences in CMR association between men and women, and in age at CMR (<60, 60-70, >70). The interaction between subgroups was evaluated using likelihood ratio tests. Similarly, likelihood ratio tests were employed to explore possible non-linear associations between CMR measurements and HCM or DCM G+ using restricted cubic splines (placing knots at the 15th, 50th, and 85th percentile).

The limited amount of missing data (**Table 1**) was imputed through Multiple Imputation by Chained Equations (MICE), creating ten datasets, and applying Rubin’s rules to combine estimates across imputed datasets. Results are presented as odds ratios (OR) or hazard ratios (HR), with 95% confidence intervals (95%CI). Unless specified differently, p-values were evaluated against a multiplicity correction threshold of 6.25×10^-3^, accounting for the eight principal components necessary to explain at least 90% of the CMR variance (**Supplementary Figure S1-S2**).

**Table 1.** Baseline characteristics of the UK Biobank participants with data on CMR measurements and whole exome sequencing.

|  | *N (%) or Mean (SD)* | *Median (Q1; Q3)* | *Missing (%)* |
| --- | --- | --- | --- |
| *n* | 40,169 | - | - |
| *Age (years)* | 63.85 (7.71) | 64.00 (58.00; 70.00) | 0 (0.00) |
| *Sex* | 18888 (47.02) | - | 0 (0.00) |
| *BMI (kg/m<sup>2</sup>)* | 26.29 (4.30) | 25.70 (23.33; 28.56) | 1419 (3.53) |
| *Hypertension* | 11397 (28.37) | - | 0 (0.00) |
| *Diabetes* | 1224 (3.05) | - | 0 (0.00) |
| *Ever smoked* | 16097 (40.07) | - | 0 (0.00) |
| *Hypercholesterolaemia* | 8571 (21.34) | - | 0 (0.00) |
| *Family history of heart disease* | 21544 (53.63) | - | 0 (0.00) |
| *European ethnicity* | 38924 (96.90) | - | 0 (0.00) |
| *LV-EDV (ml)* | 144.24 (30.53) | 140.66 (121.05; 164.10) | 1354 (3.37) |
| *LV-ESV (ml)* | 56.74 (16.49) | 54.25 (44.34; 66.89) | 1354 (3.37) |
| *LV-ESVi (ml)* | 30.41 (7.53) | 29.55 (24.96; 34.96) | 2669 (6.64) |
| *LV-SV (ml)* | 87.50 (18.60) | 85.61 (73.93; 99.08) | 1354 (3.37) |
| *LV-EF (%)* | 60.95 (6.03) | 61.03 (57.01; 65.14) | 1354 (3.37) |
| *LV mass (g)* | 91.53 (22.34) | 88.25 (73.77; 106.84) | 1354 (3.37) |
| *RV-EDV (ml)* | 154.20 (34.44) | 149.87 (127.78; 177.34) | 1450 (3.61) |
| *RV-ESV (ml)* | 63.49 (18.47) | 61.00 (49.39; 75.29) | 1450 (3.61) |
| *RV-ESVi (ml)* | 34.02 (8.29) | 33.18 (27.94; 39.22) | 2753 (6.85) |
| *RV-SV (ml)* | 90.70 (20.32) | 88.54 (75.93; 103.69) | 1450 (3.61) |
| *RV-EF (%)* | 59.10 (5.88) | 59.16 (55.31; 63.02) | 1450 (3.61) |
| *LAVi min (ml)* | 13.79 (5.94) | 13.08 (9.57; 16.99) | 7517 (18.71) |
| *LAVi max (ml)* | 38.60 (9.76) | 37.97 (31.66; 44.78) | 7613 (18.95) |
| *LA-CI (v/v)* | 0.18 (0.07) | 0.17 (0.13; 0.22) | 7397 (18.41) |
| **LA-EF (%)** | 65.18 (8.45) | 65.04 (59.79; 70.75) | 6398 (15.93) |
| **LA res (ml)** | 21.43 (7.39) | 20.91 (16.01; 26.22) | 6614 (16.47) |
| **LA pump (ml)** | 23.05 (7.11) | 22.41 (18.00; 27.43) | 6495 (16.17) |
| **RAVi min (ml)** | 22.81 (8.19) | 21.77 (17.11; 27.33) | 9014 (22.44) |
| **RAVi max (ml)** | 46.24 (12.43) | 44.62 (37.30; 53.49) | 9081 (22.61) |
| **RA-EF (%)** | 51.02 (9.23) | 50.33 (44.68; 56.57) | 7882 (19.62) |
| **MAPSE 2Ch (mm)** | 16.60 (2.98) | 16.45 (14.50; 18.56) | 870 (2.17) |
| **TAPSE 4Ch (mm)** | 13.41 (4.71) | 12.43 (10.45; 15.13) | 1621 (4.04) |

Abbreviations: CI = compliance index, CMR = cardiac magnetic resonance imaging, EDV = end-diastolic volume, EF = ejection fraction, ESV = end-systolic volume, i = body surface area indexed, LA = left atrial, LV = left ventricular, MAPSE 2Ch = mitral annular plane systolic excursion in 2-chamber view, pump = pump volume, RA = right atrial, res = reservoir volume, RV = right ventricular, SV = stroke volume, TAPSE 4Ch = tricuspid annular plane systolic excursion in 4-chamber view, Vi max = maximum indexed volume, Vi min = minimum indexed volume.

The Q-test for heterogeneity was employed to determine to what extent the identified CMR associations with any G+ differed across the individual HCM or DCM genes involved; focussing on individual genes with at least 15 carriers (*MYBPC3*, *TNNT2*, and *MYH7* for HCM, and *TTN and MYH7* for DCM). We furthermore set out to identify gene-specific CMR associations using the same approach as the main analysis considering any HCM or DCM variant per individual gene.

### Genomics validation of CMR findings

Finally, for the subset of CMR measurements associating with G+, we sought to identify genetic support of this association by identifying common genetic variants located within or around known CMP variants and determining their association with these same CMR measurements. Therefore, we leveraged CMR GWAS available in the NHGRI-EBI GWAS Catalog^16^, extracted the genome-wide significant genetic variants, and determined which variants were located in a 1 megabase pair window around known HCM or DCM genes^9–11,17–19^.

## Results

CMR measurements and WES were available for 40,169 participants free of cardiac disease at the time of imaging. This included 248 (0.62%) participants with an HCM-associated variant and 144 (0.36%) with a DCM-associated variant. The most common genes associated with HCM were *MYBPC3* (45.6%), *TNNT2* (27.4%), or *MYH7* (19.4%), and with DCM *TTN* (30.6%), *MYH7* (23.6%), and *FLNC* (10.4 %; **Supplementary Figure S3**). The median age of the participants was 64 years (interquartile range [IQR] 58; 70), 47% was male, and the participants had a median LV-EF of 61% (IQR 57; 65), and RV-EF of 59% (IQR 55; 63; **Table 1**, **Supplementary Table S3**).

### CMR associations with the onset of AF and HF

The 22 DL-derived CMR measurements were first empirically validated by determining their association with incident AF and HF, identifying thirteen measurements associated with AF and fifteen with HF (**Figure 1-2, Supplementary table S4-S5**). For example, per standard deviation increase of LV-EF, the HR was 0.69 (95%CI 0.53; 0.89) for incident AF and 0.61 (95%CI 0.54; 0.69) for incident HF. For RV-EF this was 0.81 (95%CI 0.72; 0.90) for AF and 0.81 (95%CI 0.71; 0.92) for HF, and for LA-EF this was 0.51 (95%CI 0.46; 0.57) for AF and 0.52 (95%CI 0.46; 0.59) for HF. Furthermore, we observed that larger values of LV-EDV, LV-ESV, LV-ESVi, LV mass, RV-ESV, LA-Vi, LA compliance index, LA pump volume, and RA-Vi increased the risk of developing both AF and HF. An increase in LA reservoir volume (OR 0.73, 95%CI 0.62; 0.85) and MAPSE 2Ch (OR 0.56, 95%CI 0.48; 0.65) was associated with a decreased risk of HF but not AF (**Figure 1-2, Supplementary Tables S4-S5**). The Kaplan Meier analysis is depicted in **Supplementary Figures S4-S5**.

**Figure 1.**
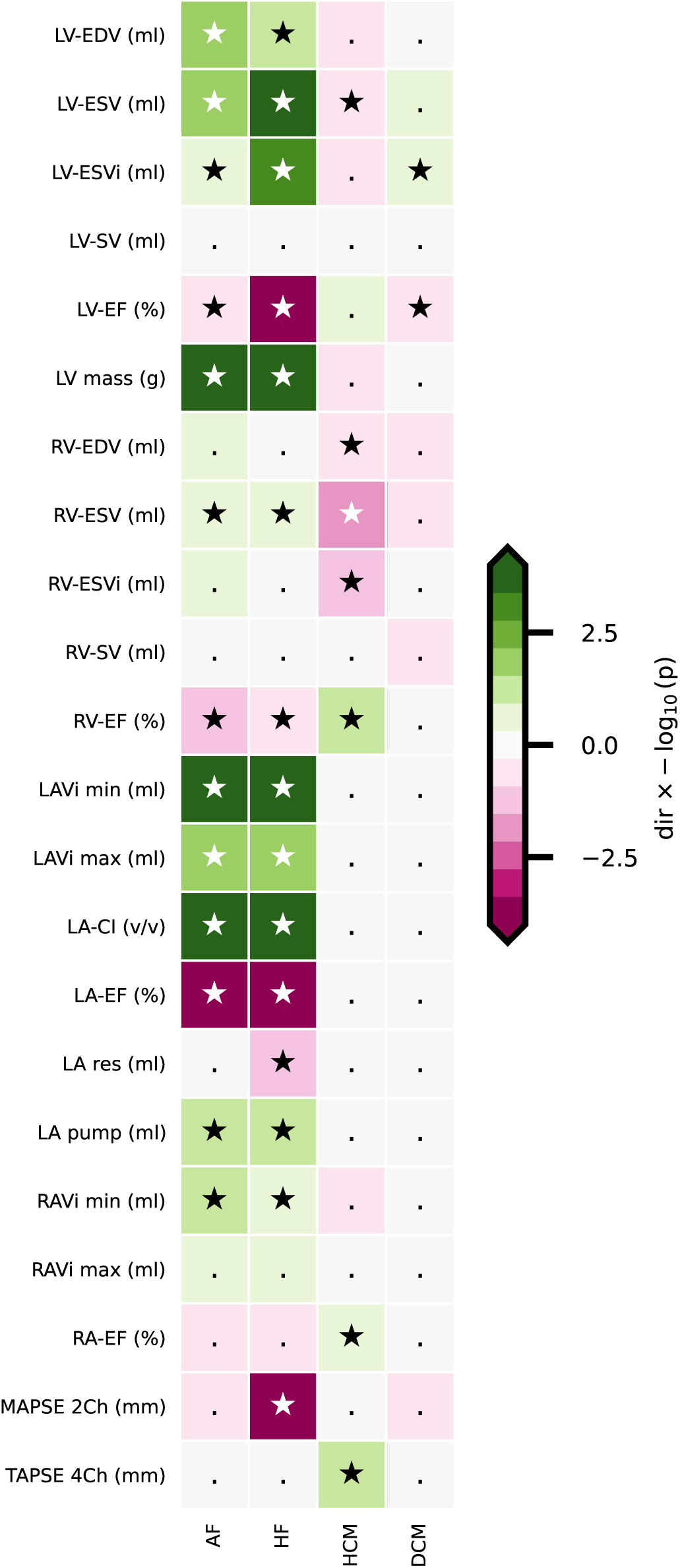
Association of CMR measurements with incident atrial fibrillation or heart failure and carriership of genetic variants associated with hypertrophic cardiomyopathy or dilated cardiomyopathy. Associations are presented as –log_10_(p-value) multiplied by the effect direction. Significant results, as defined by the Bonferroni-corrected p-value threshold of 6.25×10^-3^, are indicated with a star. Abbreviations: AF = atrial fibrillation, CI = compliance index, CMR = cardiac magnetic resonance imaging, DCM = dilated cardiomyopathy, EDV = end-diastolic volume, EF = ejection fraction, ESV = end-systolic volume, HCM = hypertrophic cardiomyopathy, HF = heart failure, i = body surface area indexed, LA = left atrial, LV = left ventricular, MAPSE 2Ch = mitral annular plane systolic excursion in 2-chamber view, pump = pump volume, RA = right atrial, res = reservoir volume, RV = right ventricular, SV = stroke volume, TAPSE 4Ch = tricuspid annular plane systolic excursion in 4-chamber view, Vi max = maximum indexed volume, Vi min = minimum indexed volume.

**Figure 2.**
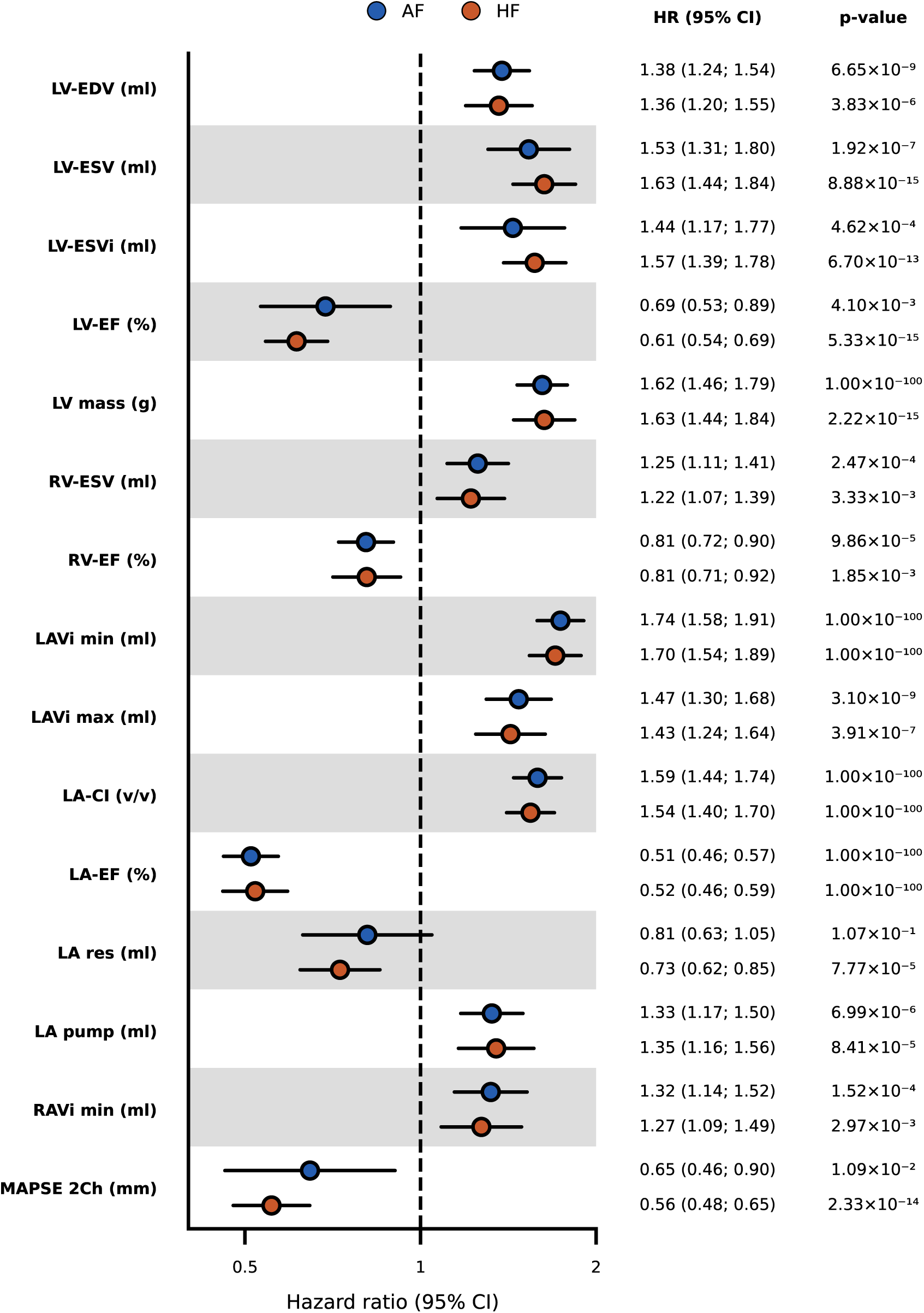
Hazard ratios for the association of CMR measurements with incident atrial fibrillation or heart failure. Effect magnitudes are presented per standard deviation increase in CMR measurements. Abbreviations: AF = atrial fibrillation, CI = compliance index, CMR = cardiac magnetic resonance imaging, EDV = end-diastolic volume, EF = ejection fraction, ESV = end-systolic volume, HF = heart failure, HR = hazard ratio, i = body surface area indexed, LA = left atrial, LV = left ventricular, MAPSE 2Ch = mitral annular plane systolic excursion in 2-chamber view, RA = right atrial, RV = right ventricular, SV = stroke volume, TAPSE 4Ch = tricuspid annular plane systolic excursion in 4-chamber view, Vi max = maximum indexed volume, 95% CI = 95% confidence interval.

### CMR associations with G+

After accounting for known cardiac risk factors, we identified five CMR measurements associated with HCM G+ and two CMR measurements associated with DCM G+ (**Figure 3**). The odds of HCM G+ increased with larger values of RV-EF (OR 1.36, 95%CI 1.19; 1.55), RA-EF (OR 1.24 95%CI 1.07; 1.43), TAPSE 4Ch (OR 1.22, 95%CI 1.11; 1.35), and decreased with RV-ESV (OR 0.62, 95%CI 0.53; 0.74), and RV-ESVi (OR 0.69, 95%CI 0.59; 0.80; **Figure 3**, **Supplementary Table S6**). The odds of DCM G+ decreased with larger values of LV-EF (OR 0.74, 95%CI 0.63; 0.87) and increased with LV-ESVi (OR 1.36, 95%CI 1.15; 1.60; **Figure 3**, **Supplementary Table S7**). Comparing these results to the CMR associations with incident AF and HF, we found that associations with DCM G+ were typically in the same direction as that of HF and AF associations. However, for HCM G+, we generally observed that CMR measurements with incident disease were in the opposite direction (**Figure 4**). For example, larger values of RV-EF increased the odds of HCM G+ but were associated with a decreased risk of AF and HF (**Figure 1**).

**Figure 3.**
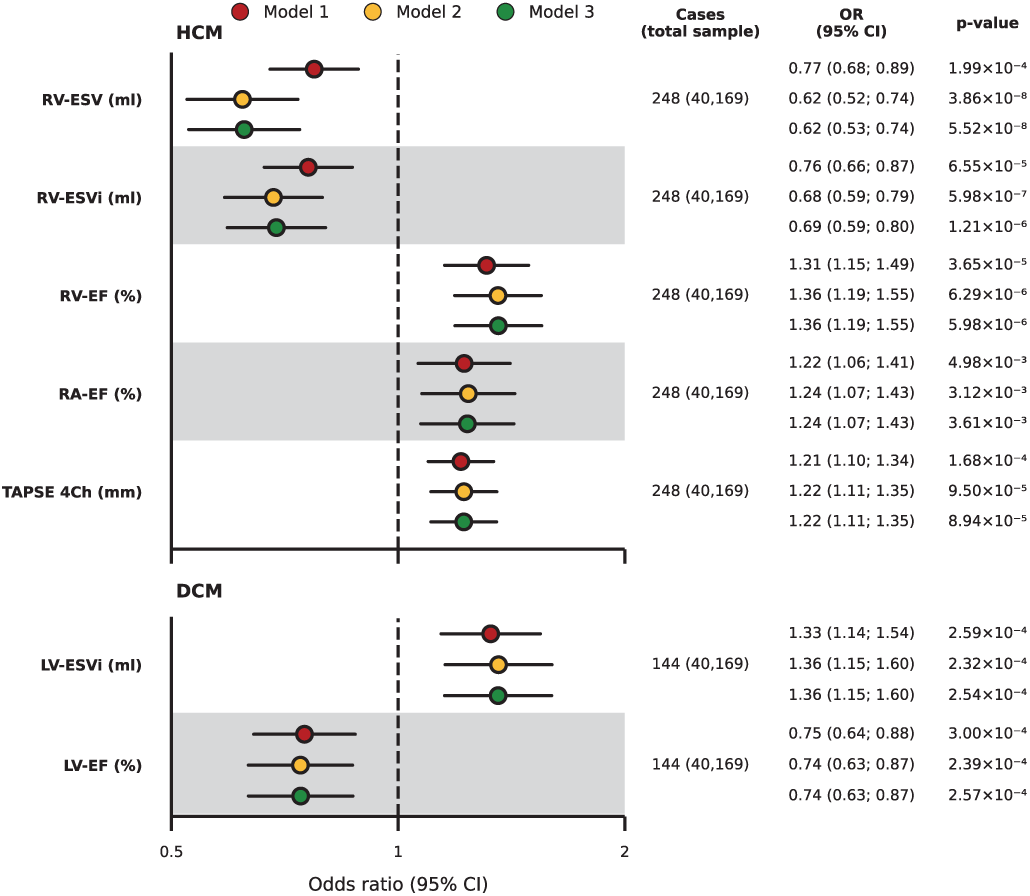
Odds ratios for the association of CMR measurements with hypertrophic cardiomyopathy or dilated cardiomyopathy G+. Model 1 is univariable, model 2 is adjusted for age and sex, model 3 is adjusted for age, sex, and the comorbidities hypertension, diabetes, hypercholesterolaemia, and smoking. Abbreviations: CMR = cardiac magnetic resonance imaging, DCM = dilated cardiomyopathy, EF = ejection fraction, ESV = end-systolic volume, HCM = hypertrophic cardiomyopathy, i = body surface area indexed, LV = left ventricular, RA = right atrial, RV = right ventricular, OR = odds ratio, TAPSE 4Ch = tricuspid annular plane systolic excursion in 4-chamber view, 95% CI = confidence interval.

**Figure 4.**
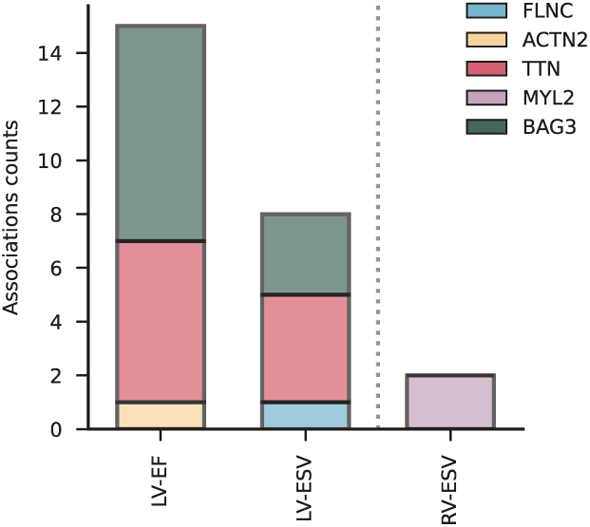
Frequency of genetic variants in cardiomyopathy-associated genes associating with CMR measurements. Genetic variants were selected within 1 megabasepair of cardiomyopathy-associated genes and searched in genome-wide association study summary statistics. Associations with dilated cardiomyopathy are depicted left of the vertical line, those with hypertrophic cardiomyopathy to the right. Abbreviations: EF = ejection fraction, ESV = end-systolic volume, LV = left ventricular, RV = right ventricular.

Sensitivity analyses did not identify substantial evidence for non-linearity, or effect modification by age or sex (**Supplementary Tables S8-S10**). Restricting the included variants to only pathogenic or likely pathogenic submissions did not alter any of the associations (**Supplementary Table S6-S7**).

### Gene specific CMR associations

We then focused on pathogenic and likely pathogenic variants in the HCM and DCM genes with at least 15 carriers (*MYBPC3*, *TNNT2*, and *MYH7* for HCM and *TTN* and *MYH7* for DCM). Subsequently, we assessed whether CMR measurements specifically associated with variants in these six individual genes. In addition to the overall associations with HCM, we found that *TNNT2* uniquely associated with MAPSE 2Ch (OR 1.41, 95%CI 1.11; 1.79), and *MYBPC3* associated with LV-EDV (OR 0.70, 95%CI 0.55; 0.90) and RV-EDV (OR 0.58,

95%CI 0.45; 0.75). The associations of LV-EF and LV-ESVi with DCM G+ showed substantial heterogeneity, which were driven by variants in *TTN* (**Supplementary Figures S6-S7**, **Supplementary Table S11-12**). For DCM, we identified specific CMR associations with *TTN* (MAPSE 2Ch OR 0.58, 95%CI 0.41; 0.81) and *MYH7* (LA pump OR 1.70, 95%CI 1.21; 2.38, and RA-EF OR 1.61, 95%CI 1.16; 2.23; **Supplementary Figures S8-S9**, **Supplementary Table S12**).

### Genomic validation of identified CMR biomarkers

To further validate our findings, we identified common genetic variants located within and around known HCM and DCM genes, that were associated with the subset of CMR measurements linked to G+. For this, we extracted genome-wide significant findings from GWAS conducted on biventricular EF and ESV, RV-EDV, RV-SV, and RA-EF. We found that fifteen genetic associations of variants near or within causal genes for DCM (*BAG3*, *TTN*, and *ACTN2*) also associated with LV-EF (**Figure 4**, **Supplementary Table S13**). Similarly, eight genetic associations near known DCM genes were observed for LV-ESV (**Figure 4**).

For CMR measurements associating with HCM G+ we were able to genetically validate the association of RV-ESV, finding two genetic associations within/near *MYL2* associating with this measurement. A similar validation step could not be conducted for TAPSE because this measurement has not yet been considered in GWAS.

## Discussion

In the current study, we empirically validated DL-based automatic analysis of 22 CMR measurements by confirming known associations with incident AF and HF. We subsequently combined these measurements with WES data to establish imaging biomarkers of the left and right ventricle and the right atrium in participants carrying variants associated with HCM or DCM. Focussing on the three most common genes associated with HCM and DCM, we identified gene-specific effects. Lastly, we provide genetic validation, confirming that common genetic variants within or around CMP-causing genes were associated with the same CMR measurements. These validated CMR measurements offer insights into the pre-clinical phenotypes of CMP, and provide potential surrogate endpoints for clinical trials evaluating novel therapeutics^20,21^.

Of the 22 CMR measurements, thirteen associated with incident AF and fifteen with HF, confirming subtle structural and functional cardiac abnormalities in these diseases. Known CMR measurements associating with AF and HF included LV-EF, LV-mass, RV-ESV, RV-EF, and LA volume. All measurements associated with AF also associated with HF in the same direction, but HF was associated with two additional CMR measurements. These results suggest that DL-based automatic analysis of CMR is a feasible modality for risk stratification in early cardiac disease.

Next, we established novel CMR measurements that associate with carriership of variants associated with HCM and DCM (G+). Primarily right heart measurements associated with HCM G+, namely RV-EF (OR 1.36, 95%CI 1.19; 1.55), RV-ESV (OR 0.62, 95%CI 0.53; 0.74), RV-ESVi (OR 0.69, 95%CI 0.59; 0.80), RA-EF (OR 1.24, 95%CI 1.07; 1.43), and TAPSE 4Ch (OR 1.22, 95%CI 1.11; 1.35). While HCM in patients is typically characterised by LV hypertrophy^2^, our findings suggest that LV hypertrophy does not associate strongly with HCM G+ in participants without cardiac disease. RV measurements have previously been found to be highly predictive of HCM progression, which associated with an increased risk of supraventricular and ventricular arrhythmias, progressive HF, and sudden cardiac death^20,21^. The role of right heart characteristics in early or pre-clinical HCM remains less characterised. Our findings may reflect adaptive changes in response to subtle diastolic LV dysfunction, a frequent early indicator of HCM that can often be detected by ultrasound, but not by CMR. This underscores the importance of investigating right heart remodelling and its origins as potential markers of subclinical HCM development. The associations with RV-EF and RV-ESV were in opposite effect directions compared to those in AF and HF, underscoring a notable divergence in pre-clinical cardiac phenotypes. For example, where a standard deviation increase in RV-EF decreased risk of incident AF and HF, it increased the odds of HCM G+. Similar directional discordance was observed in GWAS of HCM and DCM, where the genetic correlation showed a near ubiquitous opposing direction of association.

For example, the genetic correlation between LV-ESV and HCM was –0.31 and that with DCM was 0.46^18^. This directional discordance between HCM G+ and disease onset supports the potential occurrence of structural changes in the right heart, which lead to functional gains before resulting in clinical burden. Furthermore, only a subset of the CMR measurements associated with CMP G+ were also associated with AF and HF onset. As such, carriership of these variants does not simply reflect early signs of AF or HF, but instead represents a unique pre-clinical phenotype otherwise overlooked.

LV-EF (OR 0.74, 95%CI 0.63; 0.87) and LV-ESVi (OR 1.36, 95%CI 1.15; 1.60) were associated with DCM G+ in the same direction as the associations with AF and HF. LV-EF is a strong predictive marker for HF in DCM and together with LV-ESVi the most frequently used imaging marker for DCM diagnosis and monitoring^22^. We observed considerable heterogeneity in the three most common genes associated with DCM and showed that *TTN* drives the association with LV-EF. These results indicate that the pre-clinical manifestations of variants in specific genes are heterogeneous, and it may be beneficial to consider them as distinct entities.

CMR measurements not only enhance disease diagnosis and risk stratification, but also serve as surrogate markers for therapeutic efficacy^23^. The myosin inhibitor mavacamten, for example, was approved for use in HCM partly based on its effect on LV outflow tract gradient^24,25^. Our research expands on this by identifying early phenotype imaging biomarkers relevant to identify individuals carrying variants associated with CMP at a higher risk of developing the disease. While several imaging biomarkers are commonly utilized, primarily those related to LV function, dimension, and mass, we demonstrated the importance of considering RV measurements for HCM. Finally, our findings indicate significant phenotypic differences in CMR associations and pathogenic variants in DCM genes. We observed different associations for LV-EF with *TTN* compared to the other genes, which may reflect distinct underlying mechanisms for pathogenesis depending on the specific genetic variant.

This study has a number of potential limitations. We were unable to confirm whether the identified CMR measurements also associate with the development of cardiac disease in G+, because of the limited sample size. This may be especially relevant for people carrying an HCM variant, because the association with RV and RA measurements is less well established. Similarly, in addition to the identified imaging biomarkers for pre-clinical HCM and DCM phenotypes, larger sample size research is needed to explore the relevance of non-CMR parameters such as general patient characteristics or electrocardiography measurements. Combining these factors will lead to a holistic risk prediction model and this study contributes to such an approach. Additionally, our dataset predominantly comprises individuals of European ethnicity, potentially limiting the generalizability of our findings to more diverse populations.

### Conclusions

In conclusion, right heart measurements are associated with HCM G+, while LV measurements associate with DCM G+ in individuals without established cardiac disease. The identified RV and RA measurements associating with HCM G+ reflect enhanced cardiac function, potentially indicating transient compensatory mechanisms. The observed variability in CMR associations with DCM genes suggests that early cardiac phenotype differs by individual genes, which is in line with current understanding of clinical manifestations being gene specific.

## Data Availability

All scripts and aggregated data are available online at <URL available upon publication>.

## Acknowledgements

This research has been conducted using the UK Biobank Resource under application numbers 12113 and 24711. The authors are grateful to the UK Biobank participants.

## Non-standard abbreviations and acronyms

AF: atrial fibrillation
CMP: cardiomyopathy
CMR: cardiac magnetic resonance imaging
DL: deep learning
DCM: dilated cardiomyopathy
EDV: end-diastolic volume
EF: ejection fraction
ESV: end-systolic volume
GWAS: genome-wide association study
G+: individuals carrying disease-associated variants
HCM: hypertrophic cardiomyopathy
HF: heart failure
HR: hazard ratio
i: indexed
IQR: interquartile range
LA: left atrial
LV: left ventricular
MAPSE: mitral annular plane systolic excursion
OR: odds ratio
Pump: pump volume
RA: right atrial
Res: reservoir volume
RV: right ventricular
SV: stroke volume
TAPSE: tricuspid annular plane systolic excursion
V max: maximal volume
V min: minimal volume
WES: whole exome sequencing
2Ch: 2-chamber
4Ch: 4-chamber
95%CI: 95% confidence interval

## Author contributions

AFS, MvV, and PMC designed the study. MvV and PMC performed the analyses and drafted the manuscript. AFS, CPA, MW, RK, FWA provided critical input on the analysis, as well as the drafted manuscript. All authors read and approved the final manuscript.

## Code and data availability

Analyses were conducted using python 3.11. For full code availability see repository: <URL AVAILABLE UPON PUBLICATION>. The data used for this can be applied for with the UK biobank.

## References

1. Arbelo E, Protonotarios A, Gimeno JR, Arbustini E, Barriales-Villa R, Basso C, Bezzina CR, Biagini E, Blom NA, de Boer RA, et al. 2023 ESC Guidelines for the management of cardiomyopathies: Developed by the task force on the management of cardiomyopathies of the European Society of Cardiology (ESC). Eur. Heart J. 2023;44:3503–3626.

2. Ingles J, Burns C, Bagnall RD, Lam L, Yeates L, Sarina T, Puranik R, Briffa T, Atherton JJ, Driscoll T, et al. Nonfamilial hypertrophic cardiomyopathy: prevalence, natural history, and clinical implications. Circ. Cardiovasc. Genet. 2017;10:e001620.

3. Hershberger RE, Morales A, Siegfried JD. Clinical and genetic issues in dilated cardiomyopathy: a review for genetics professionals. Genet. Med. 2010;12:655–667.

4. Hershberger RE, Hedges DJ, Morales A. Dilated cardiomyopathy: the complexity of a diverse genetic architecture. Nat. Rev. Cardiol. 2013;10:531–547.

5. Grünig E, Tasman JA, Kücherer H, Franz W, Kübler W, Katus HA. Frequency and phenotypes of familial dilated cardiomyopathy. J. Am. Coll. Cardiol. 1998;31:186–194.

6. Petretta M, Pirozzi F, Sasso L, Paglia A, Bonaduce D. Review and metaanalysis of the frequency of familial dilated cardiomyopathy. Am. J. Cardiol. 2011;108:1171–1176.

7. Bourfiss M, van Vugt M, Alasiri AI, Ruijsink B, van Setten J, Schmidt AF, Dooijes D, Puyol-Antón E, Velthuis BK, van Tintelen JP, et al. Prevalence and Disease Expression of Pathogenic and Likely Pathogenic Variants Associated With Inherited Cardiomyopathies in the General Population. Circ. Genomic Precis. Med. 2022;15:e003704.

8. Ruijsink B, Puyol-Antón E, Oksuz I, Sinclair M, Bai W, Schnabel JA, Razavi R, King AP. Fully automated, quality-controlled cardiac analysis from CMR: validation and large-scale application to characterize cardiac function. *Cardiovasc*. Imaging. 2020;13:684– 695.

9. Aung N, Lopes LR, van Duijvenboden S, Harper AR, Goel A, Grace C, Ho CY, Weintraub WS, Kramer CM, Neubauer S, et al. Genome-wide analysis of left ventricular maximum wall thickness in the UK biobank cohort reveals a shared genetic background with hypertrophic cardiomyopathy. Circ. Genomic Precis. Med. 2023;16:e003716.

10. Pirruccello JP, Bick A, Wang M, Chaffin M, Friedman S, Yao J, Guo X, Venkatesh BA, Taylor KD, Post WS, et al. Analysis of cardiac magnetic resonance imaging in 36,000 individuals yields genetic insights into dilated cardiomyopathy. Nat. Commun. 2020;11:2254.

11. Schmidt AF, Bourfiss M, Alasiri A, Puyol-Anton E, Chopade S, van Vugt M, van der Laan SW, Gross C, Clarkson C, Henry A, et al. Druggable proteins influencing cardiac structure and function: Implications for heart failure therapies and cancer cardiotoxicity. Sci. Adv. 2023;9:eadd4984.

12. Strande NT, Riggs ER, Buchanan AH, Ceyhan-Birsoy O, DiStefano M, Dwight SS, Goldstein J, Ghosh R, Seifert BA, Sneddon TP, et al. Evaluating the clinical validity of gene-disease associations: an evidence-based framework developed by the clinical genome resource. Am. J. Hum. Genet. 2017;100:895–906.

13. Ingles J, Goldstein J, Thaxton C, Caleshu C, Corty EW, Crowley SB, Dougherty K, Harrison SM, McGlaughon J, Milko LV, et al. Evaluating the clinical validity of hypertrophic cardiomyopathy genes. Circ. Genomic Precis. Med. 2019;12:e002460.

14. Jordan E, Peterson L, Ai T, Asatryan B, Bronicki L, Brown E, Celeghin R, Edwards M, Fan J, Ingles J, et al. Evidence-based assessment of genes in dilated cardiomyopathy. Circulation. 2021;144:7–19.

15. Petersen SE, Matthews PM, Francis JM, Robson MD, Zemrak F, Boubertakh R, Young AA, Hudson S, Weale P, Garratt S, et al. UK Biobank’s cardiovascular magnetic resonance protocol. J. Cardiovasc. Magn. Reson. 2015;18:1–7.

16. Sollis E, Mosaku A, Abid A, Buniello A, Cerezo M, Gil L, Groza T, Güneş O, Hall P, Hayhurst J, et al. The NHGRI-EBI GWAS Catalog: knowledgebase and deposition resource. Nucleic Acids Res. 2023;51:D977–D985.

17. Choquet H, Thai KK, Jiang C, Ranatunga DK, Hoffmann TJ, Go AS, Lindsay AC, Ehm MG, Waterworth DM, Risch N, et al. Meta-analysis of 26 638 individuals identifies two genetic loci associated with left ventricular ejection fraction. Circ. Genomic Precis. Med. 2020;13:e002804.

18. Tadros R, Francis C, Xu X, Vermeer AMC, Harper AR, Huurman R, Kelu Bisabu K, Walsh R, Hoorntje ET, te Rijdt WP, et al. Shared genetic pathways contribute to risk of hypertrophic and dilated cardiomyopathies with opposite directions of effect. Nat. Genet. 2021;53:128–134.

19. Forrest IS, Rocheleau G, Bafna S, Argulian E, Narula J, Natarajan P, Do R. Genetic and phenotypic profiling of supranormal ejection fraction reveals decreased survival and underdiagnosed heart failure. Eur. J. Heart Fail. 2022;24:2118–2127.

20. Roşca M, Călin A, Beladan CC, Enache R, Mateescu AD, Gurzun M-M, Varga P, Băicuş C, Coman IM, Jurcuţ R, et al. Right ventricular remodeling, its correlates, and its clinical impact in hypertrophic cardiomyopathy. J. Am. Soc. Echocardiogr. 2015;28:1329–1338.

21. Mahmod M, Raman B, Chan K, Sivalokanathan S, Smillie RW, Abd Samat AH, Ariga R, Dass S, Ormondroyd E, Watkins H, et al. Right ventricular function declines prior to left ventricular ejection fraction in hypertrophic cardiomyopathy. J. Cardiovasc. Magn. Reson. 2022;24:36.

22. Becker MA, Cornel JH, Van de Ven PM, van Rossum AC, Allaart CP, Germans T. The prognostic value of late gadolinium-enhanced cardiac magnetic resonance imaging in nonischemic dilated cardiomyopathy: a review and meta-analysis. JACC Cardiovasc. Imaging. 2018;11:1274–1284.

23. Benz DC, Gräni C, Antiochos P, Heydari B, Gissler MC, Ge Y, Cuddy SA, Dorbala S, Kwong RY. Cardiac magnetic resonance biomarkers as surrogate endpoints in cardiovascular trials for myocardial diseases. Eur. Heart J. 2023;44:4738–4747.

24. Desai MY, Owens A, Wolski K, Geske JB, Saberi S, Wang A, Sherrid M, Cremer PC, Lakdawala NK, Tower-Rader A, et al. Mavacamten in patients with hypertrophic cardiomyopathy referred for septal reduction: week 56 results from the VALOR-HCM randomized clinical trial. JAMA Cardiol. 2023;8:968–977.

25. Olivotto I, Oreziak A, Barriales-Villa R, Abraham TP, Masri A, Garcia-Pavia P, Saberi S, Lakdawala NK, Wheeler MT, Owens A, et al. Mavacamten for treatment of symptomatic obstructive hypertrophic cardiomyopathy (EXPLORER-HCM): a randomised, double-blind, placebo-controlled, phase 3 trial. The Lancet. 2020;396:759–769.

