## Supplementary Materials for "Cardiac magnetic resonance markers of pre-clinical hypertrophic and dilated cardiomyopathy in genetic variant carriers"

**Content**

Methods……………………………………………………………………………………………..2
Figure legends…………………………………………..………………………………3
Figure S1..…………………………………………………………………………………..6
Figure S2..…………………………………………………………………………………..7
Figure S3..…………………………………………………………………………………..8
Figure S4..…………………………………………………………………………………..9
Figure S5..………………………………………………………………………………….10
Figure S6..………………………………………………………………………………….11
Figure S7..………………………………………………………………………………….12
Figure S8..………………………………………………………………………………….13
Figure S9..………………………………………………………………………………….14

### Methods

*Data engineering strategy*

We developed a de novo data engineering pipeline for extracting outcome data from the UK Biobank called “PhenotypeConstructor”. Its core functionality revolves around processing a range of disease definitions to extract the earliest diagnosis dates for various conditions. This includes interpreting ICD diagnosis codes, self-reported diagnoses, doctor-confirmed conditions, as well as medication details, and family health history. The phenotypes, corresponding UK Biobank field names, and codes considered in this study are described in **Supplementary Table S2**.

### Figure legends

**Figure S1. Loadings of the eight principal components explaining 90% of the variance in the CMR measurements.**

Abbreviations: EF = ejection fraction, EDV = end-diastolic volume, ESV = end-systolic volume, i = indexed, LA = left atrial, LV = left ventricular, MAPSE = mitral annular plane systolic excursion, PC = principal component, RA = right atrial, RV = right ventricular, SV = stroke volume, TAPSE = tricuspid annular plane systolic excursion, V = volume, 2Ch = in 2-chamber view.

**Figure S2. Cumulative variance explained by the eight principal components explaining 90% of the CMR measurements.**

**Figure S3. Distribution of UK biobank participants carrying pathogenic and likely pathogenic variants in cardiomyopathy-associated genes.**

**Figure S4. Survival of incident atrial fibrillation based on CMR measurements.**

The curves estimate cumulative incidence for atrial fibrillation categorising the CMR measurements into two groups: an 85% 'reference' group and a 15% 'risk increasing' group, based on either the $15^{th}$ or $85^{th}$ percentile of CMR measurements as the cut-off point. Each plot is annotated with the hazard ratio derived from univariable Cox regression, along with the corresponding confidence interval and p-value.

Abbreviations: EF = ejection fraction, EDV = end-diastolic volume, ESV = end-systolic volume, i = indexed, LA = left atrial, LV = left ventricular, RA = right atrial, RV = right ventricular, SV = stroke volume, TAPSE = tricuspid annular plane systolic excursion, V = volume, 4Ch = in 4-chamber view.

**Figure S5. Survival of incident heart failure based on CMR measurements.**

The curves estimate cumulative incidence for heart failure categorising the CMR measurements into two groups: an 85% 'reference' group and a 15% 'risk increasing' group, based on either the 15th or 85th percentile of CMR measurements as the cut-off point. Each plot is annotated with the hazard ratio derived from univariable Cox regression, along with the corresponding confidence interval and p-value.

Abbreviations: EF = ejection fraction, EDV = end-diastolic volume, ESV = end-systolic volume, i = indexed, LA = left atrial, LV = left ventricular, RA = right atrial, RV = right ventricular, SV = stroke volume, TAPSE = tricuspid annular plane systolic excursion, V = volume, 4Ch = in 4-chamber view.

**Figure S6. Association of CMR measurements with HCM G+ and the three most common HCM genes.**

Associations are presented as $-\log_{10}$(p-value) multiplied by the effect direction. Results with a p-value smaller than $6.25\times10^{-3}$ are indicated by a star and smaller than 0.05 are indicated with a diamond. Heterogeneity p-values did not reach statistical significance for any of the CMR measurements.

Abbreviations: CMR = cardiac magnetic resonance imaging, EF = ejection fraction, ESV = end-systolic volume, HCM = hypertrophic cardiomyopathy, i = indexed, RA = right atrial, RV = right ventricular, TAPSE = tricuspid annular plane systolic excursion, V = volume, 4Ch = in 4-chamber view.

**Figure S7. Association of CMR measurements with DCM G+ and the three most common DCM genes.**

Associations are presented as $-\log_{10}$(p-value) multiplied by the effect direction. Results with a p-value smaller than $6.25\times10^{-3}$ are indicated by a star and smaller than 0.05 are indicated with a diamond. Heterogeneity p-values reached statistical significance for all of the CMR measurements.

Abbreviations: CMR = cardiac magnetic resonance imaging, DCM = dilated cardiomyopathy, EF = ejection fraction, ESV = end-systolic volume, i = indexed, LV = left ventricular.

**Figure S8. Association of CMR measurements with the most common HCM genes.** None of the associations of CMR measurements with *MYH7* reached statistical significance. Abbreviations: CMR = cardiac magnetic resonance imaging, EF = ejection fraction, EDV = end-diastolic volume, ESV = end-systolic volume, HCM = hypertrophic cardiomyopathy, i = indexed, LV = left ventricular, MAPSE = mitral annular plane systolic excursion, OR = odds ratio, RV = right ventricular, TAPSE = tricuspid annular plane systolic excursion, 4Ch = in 4-chamber view, 95%CI = 95% confidence interval.

**Figure S9. Association of CMR measurements with the most common DCM genes.** None of the associations of CMR measurements with *FLNC* reached statistical significance. Abbreviations: CMR = cardiac magnetic resonance imaging, DCM = dilated cardiomyopathy, EF = ejection fraction, ESV = end-systolic volume, i = indexed, LV = left ventricular, MAPSE = mitral annular plane systolic excursion, OR = odds ratio, pump = pump volume, 2Ch = in 2-chamber view, 95%CI = 95% confidence interval.

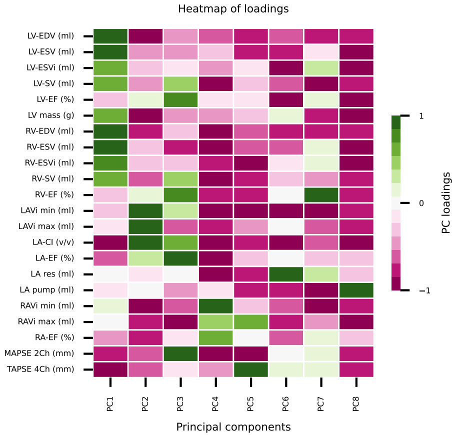

Heatmap of loadings
LV-EDV (ml)
LV-ESV (ml)
LV-ESVi (ml)
LV-SV (ml)
LV-EF (%)
LV mass (g)
RV-EDV (ml)
RV-ESV (ml)
RV-ESVi (ml)
RV-SV (ml)
RV-EF (%)
LAVi min (ml)
LAVi max (ml)
LA-CI (v/v)
LA-EF (%)
LA res (ml)
LA pump (ml)
RAVi min (ml)
RAVi max (ml)
RA-EF (%)
MAPSE 2Ch (mm)
TAPSE 4Ch (mm)
PC1
PC2
PC3
PC4
PC5
PC6
PC7
PC8
Principal components
1
0
−1
PC loadings

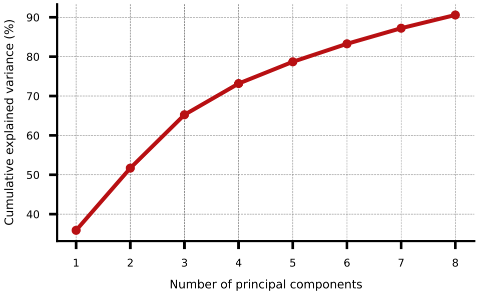
Cumulative explained variance (%)
90
80
70
60
50
40
1
2
3
4
5
6
7
8
Number of principal components

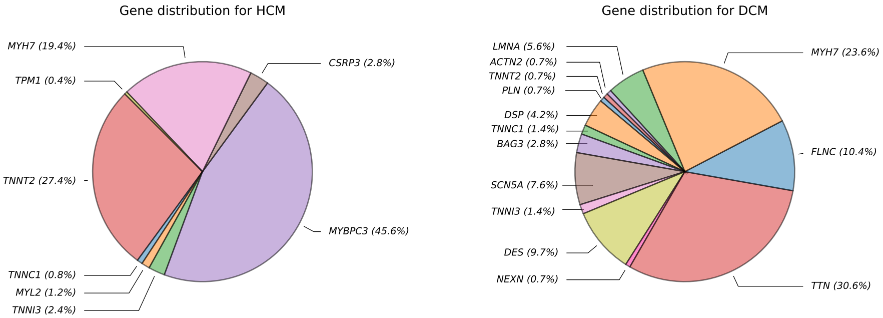
Gene distribution for HCM
MYH7 (19.4%)
TPM1 (0.4%)
TNNT2 (27.4%)
TNNC1 (0.8%)
MYL2 (1.2%)
TNNI3 (2.4%)
CSRP3 (2.8%)
MYBPC3 (45.6%)
Gene distribution for DCM
LMNA (5.6%)
ACTN2 (0.7%)
TNNT2 (0.7%)
PLN (0.7%)
DSP (4.2%)
TNNC1 (1.4%)
BAG3 (2.8%)
SCN5A (7.6%)
TNNI3 (1.4%)
DES (9.7%)
NEXN (0.7%)
MYH7 (23.6%)
FLNC (10.4%)
TTN (30.6%)

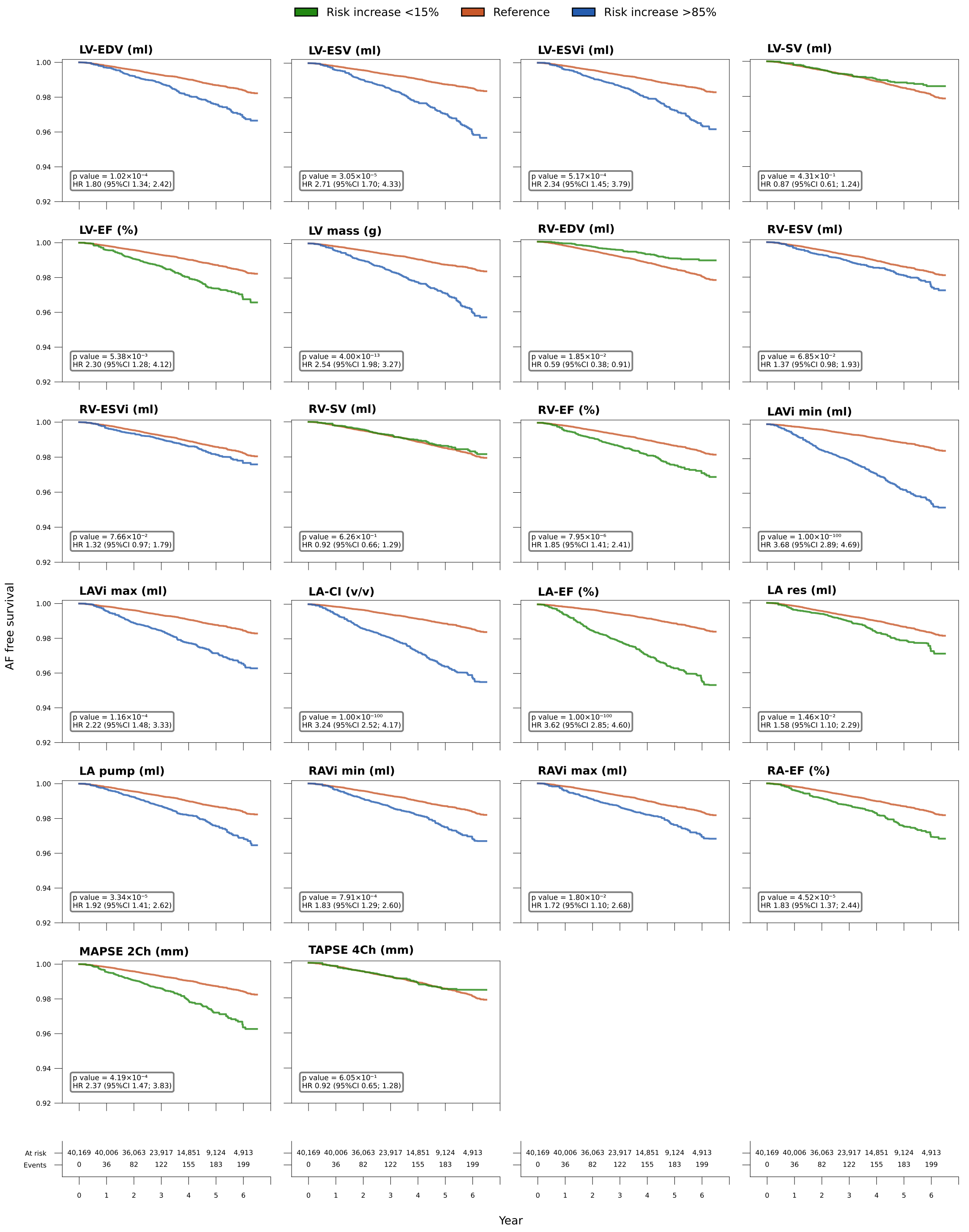

Risk increase <15%
Reference
Risk increase >85%
AF free survival
LV-EDV (ml)
p value = $1.02\times10^{-4}$
HR 1.80 (95%CI 1.34; 2.42)
LV-ESV (ml)
p value = $3.05\times10^{-5}$
HR 2.71 (95%CI 1.70; 4.33)
LV-ESVi (ml)
p value = $5.17\times10^{-4}$
HR 2.34 (95%CI 1.45; 3.79)
LV-SV (ml)
p value = $4.31\times10^{-1}$
HR 0.87 (95%CI 0.61; 1.24)
LV-EF (%)
p value = $5.38\times10^{-3}$
HR 2.30 (95%CI 1.28; 4.12)
LV mass (g)
p value = $4.00\times10^{-13}$
HR 2.54 (95%CI 1.98; 3.27)
RV-EDV (ml)
p value = $1.85\times10^{-2}$
HR 0.59 (95%CI 0.38; 0.91)
RV-ESV (ml)
p value = $6.85\times10^{-2}$
HR 1.37 (95%CI 0.98; 1.93)
RV-ESVi (ml)
p value = $7.66\times10^{-2}$
HR 1.32 (95%CI 0.97; 1.79)
RV-SV (ml)
p value = $6.26\times10^{-1}$
HR 0.92 (95%CI 0.66; 1.29)
RV-EF (%)
p value = $7.95\times10^{-6}$
HR 1.85 (95%CI 1.41; 2.41)
LAVi min (ml)
p value = $1.00\times10^{-100}$
HR 3.68 (95%CI 2.89; 4.69)
LAVi max (ml)
p value = $1.16\times10^{-4}$
HR 2.22 (95%CI 1.48; 3.33)
LA-CI (v/v)
p value = $1.00\times10^{-100}$
HR 3.24 (95%CI 2.52; 4.17)
LA-EF (%)
p value = $1.00\times10^{-100}$
HR 3.62 (95%CI 2.85; 4.60)
LA res (ml)
p value = $1.46\times10^{-2}$
HR 1.58 (95%CI 1.10; 2.29)
LA pump (ml)
p value = $3.34\times10^{-5}$
HR 1.92 (95%CI 1.41; 2.62)
RAVi min (ml)
p value = $7.91\times10^{-4}$
HR 1.83 (95%CI 1.29; 2.60)
RAVi max (ml)
p value = $1.80\times10^{-2}$
HR 1.72 (95%CI 1.10; 2.68)
RA-EF (%)
p value = $4.52\times10^{-5}$
HR 1.83 (95%CI 1.37; 2.44)
MAPSE 2Ch (mm)
p value = $4.19\times10^{-4}$
HR 2.37 (95%CI 1.47; 3.83)
TAPSE 4Ch (mm)
p value = $6.05\times10^{-1}$
HR 0.92 (95%CI 0.65; 1.28)
1.00
0.98
0.96
0.94
0.92
At risk 40,169 40,006 36,063 23,917 14,851 9,124 4,913
Events 0 36 82 122 155 183 199
0 1 2 3 4 5 6
Year

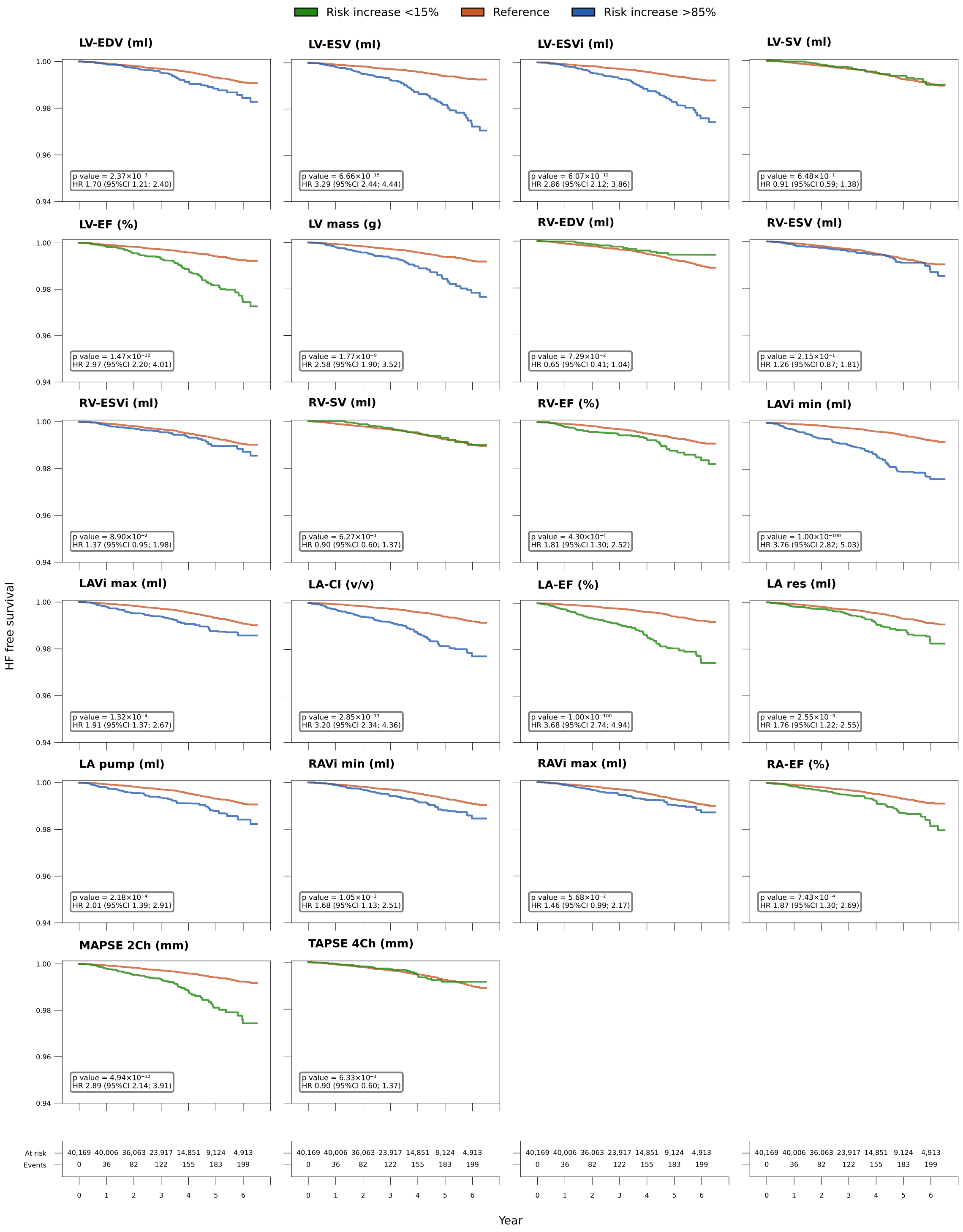

Risk increase <15%
Reference
Risk increase >85%
LV-EDV (ml)
p value = $2.37\times10^{-3}$
HR 1.70 (95%CI 1.21; 2.40)
LV-ESV (ml)
p value = $6.66\times10^{-15}$
HR 3.29 (95%CI 2.44; 4.44)
LV-ESVi (ml)
p value = $6.07\times10^{-12}$
HR 2.86 (95%CI 2.12; 3.86)
LV-SV (ml)
p value = $6.48\times10^{-1}$
HR 0.91 (95%CI 0.59; 1.38)
LV-EF (%)
p value = $1.47\times10^{-12}$
HR 2.97 (95%CI 2.20; 4.01)
LV mass (g)
p value = $1.77\times10^{-9}$
HR 2.58 (95%CI 1.90; 3.52)
RV-EDV (ml)
p value = $7.29\times10^{-2}$
HR 0.65 (95%CI 0.41; 1.04)
RV-ESV (ml)
p value = $2.15\times10^{-1}$
HR 1.26 (95%CI 0.87; 1.81)
RV-ESVi (ml)
p value = $8.90\times10^{-2}$
HR 1.37 (95%CI 0.95; 1.98)
RV-SV (ml)
p value = $6.27\times10^{-1}$
HR 0.90 (95%CI 0.60; 1.37)
RV-EF (%)
p value = $4.30\times10^{-4}$
HR 1.81 (95%CI 1.30; 2.52)
LAVi min (ml)
p value = $1.00\times10^{-100}$
HR 3.76 (95%CI 2.82; 5.03)
LAVi max (ml)
p value = $1.32\times10^{-4}$
HR 1.91 (95%CI 1.37; 2.67)
LA-CI (v/v)
p value = $2.85\times10^{-13}$
HR 3.20 (95%CI 2.34; 4.36)
LA-EF (%)
p value = $1.00\times10^{-100}$
HR 3.68 (95%CI 2.74; 4.94)
LA res (ml)
p value = $2.55\times10^{-3}$
HR 1.76 (95%CI 1.22; 2.55)
LA pump (ml)
p value = $2.18\times10^{-4}$
HR 2.01 (95%CI 1.39; 2.91)
RAVi min (ml)
p value = $1.05\times10^{-2}$
HR 1.68 (95%CI 1.13; 2.51)
RAVi max (ml)
p value = $5.68\times10^{-2}$
HR 1.46 (95%CI 0.99; 2.17)
RA-EF (%)
p value = $7.43\times10^{-4}$
HR 1.87 (95%CI 1.30; 2.69)
MAPSE 2Ch (mm)
p value = $4.94\times10^{-12}$
HR 2.89 (95%CI 2.14; 3.91)
TAPSE 4Ch (mm)
p value = $6.33\times10^{-1}$
HR 0.90 (95%CI 0.60; 1.37)
HF free survival
1.00
0.98
0.96
0.94
At risk 40,169 40,006 36,063 23,917 14,851 9,124 4,913
Events 0 36 82 122 155 183 199
0 1 2 3 4 5 6
Year

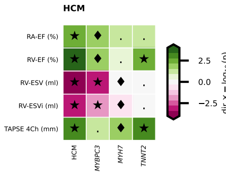

HCM
RA-EF (%)
RV-EF (%)
RV-ESV (ml)
RV-ESVi (ml)
TAPSE 4Ch (mm)
HCM
MYBPC3
MYH7
TNNT2
2.5
0.0
−2.5
dir × −log10 (p)

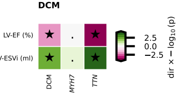

DCM
LV-EF (%)
LV-ESVi (ml)
DCM
MYH7
TTN
2.5
0.0
−2.5
dir × − log10 (p)

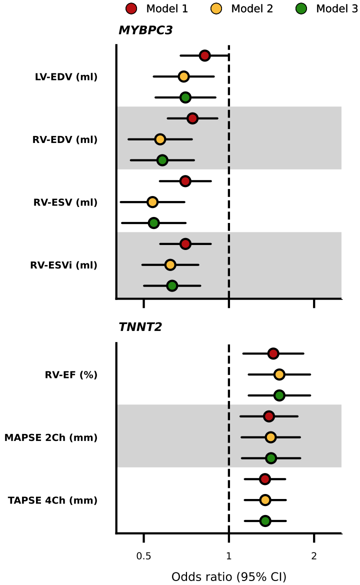


| Gene | Trait | Model | Cases (total sample) | OR (95% CI) | p-value |
|---|---|---|---|---|---|
| *MYBPC3* | LV-EDV (ml) | Model 1 | 113 (40,169) | 0.82 (0.68; 1.00) | $5.02\times10^{-2}$ |
| | | Model 2 | | 0.69 (0.54; 0.88) | $3.17\times10^{-3}$ |
| | | Model 3 | | 0.70 (0.55; 0.90) | $4.44\times10^{-3}$ |
| | RV-EDV (ml) | Model 1 | 113 (40,169) | 0.74 (0.61; 0.91) | $4.01\times10^{-3}$ |
| | | Model 2 | | 0.57 (0.44; 0.74) | $2.06\times10^{-5}$ |
| | | Model 3 | | 0.58 (0.45; 0.75) | $3.53\times10^{-5}$ |
| | RV-ESV (ml) | Model 1 | 113 (40,169) | 0.70 (0.57; 0.86) | $8.11\times10^{-4}$ |
| | | Model 2 | | 0.54 (0.41; 0.70) | $2.49\times10^{-6}$ |
| | | Model 3 | | 0.54 (0.42; 0.70) | $3.51\times10^{-6}$ |
| | RV-ESVi (ml) | Model 1 | 113 (40,169) | 0.70 (0.57; 0.86) | $7.51\times10^{-4}$ |
| | | Model 2 | | 0.62 (0.49; 0.78) | $4.07\times10^{-5}$ |
| | | Model 3 | | 0.63 (0.50; 0.79) | $7.36\times10^{-5}$ |
| *TNNT2* | RV-EF (%) | Model 1 | 68 (40,169) | 1.44 (1.12; 1.83) | $3.72\times10^{-3}$ |
| | | Model 2 | | 1.51 (1.18; 1.94) | $1.25\times10^{-3}$ |
| | | Model 3 | | 1.51 (1.17; 1.94) | $1.27\times10^{-3}$ |
| | MAPSE 2Ch (mm) | Model 1 | 68 (40,169) | 1.39 (1.10; 1.75) | $5.84\times10^{-3}$ |
| | | Model 2 | | 1.41 (1.11; 1.78) | $4.94\times10^{-3}$ |
| | | Model 3 | | 1.41 (1.11; 1.79) | $4.59\times10^{-3}$ |
| | TAPSE 4Ch (mm) | Model 1 | 68 (40,169) | 1.34 (1.14; 1.58) | $4.58\times10^{-4}$ |
| | | Model 2 | | 1.34 (1.14; 1.59) | $5.15\times10^{-4}$ |
| | | Model 3 | | 1.34 (1.14; 1.59) | $4.88\times10^{-4}$ |

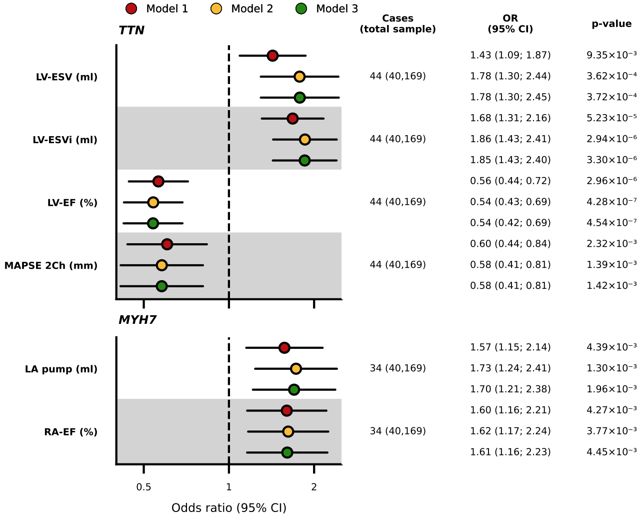
Model 1
Model 2
Model 3
TTN
Cases (total sample)
OR (95% CI)
p-value
LV-ESV (ml)
44 (40,169)
1.43 (1.09; 1.87)
9.35×10⁻³
1.78 (1.30; 2.44)
3.62×10⁻⁴
1.78 (1.30; 2.45)
3.72×10⁻⁴
LV-ESVi (ml)
44 (40,169)
1.68 (1.31; 2.16)
5.23×10⁻⁵
1.86 (1.43; 2.41)
2.94×10⁻⁶
1.85 (1.43; 2.40)
3.30×10⁻⁶
LV-EF (%)
44 (40,169)
0.56 (0.44; 0.72)
2.96×10⁻⁶
0.54 (0.43; 0.69)
4.28×10⁻⁷
0.54 (0.42; 0.69)
4.54×10⁻⁷
MAPSE 2Ch (mm)
44 (40,169)
0.60 (0.44; 0.84)
2.32×10⁻³
0.58 (0.41; 0.81)
1.39×10⁻³
0.58 (0.41; 0.81)
1.42×10⁻³
MYH7
LA pump (ml)
34 (40,169)
1.57 (1.15; 2.14)
4.39×10⁻³
1.73 (1.24; 2.41)
1.30×10⁻³
1.70 (1.21; 2.38)
1.96×10⁻³
RA-EF (%)
34 (40,169)
1.60 (1.16; 2.21)
4.27×10⁻³
1.62 (1.17; 2.24)
3.77×10⁻³
1.61 (1.16; 2.23)
4.45×10⁻³
0.5
1
2
Odds ratio (95% CI)
